# Validation of a Deep Learning Model to aid in COVID-19 Detection from Digital Chest Radiographs

**DOI:** 10.1101/2022.06.02.22275895

**Authors:** Pranav Ajmera, Amit Kharat, Sanjay Khaladkar, Tanveer Gupte, Richa Pant, Viraj Kulkarni, Vinay Duddalwar, Deepak Patkar, Mona Bhatia, Purnachandra Lamghare

## Abstract

**Introduction:** Using artificial intelligence in imaging practice helps ensure study list reprioritization, prompt attention to urgent studies, and reduces the reporting turn-around time.

**Purpose:** We tested a deep learning-based artificial intelligence model that can detect COVID-19 pneumonia patterns from digital chest radiographs.

**Material and Methods:** The deep learning model was built using the enhanced U-Net architecture with Spatial Attention Gate and Xception Encoder. The model was named DxCOVID and was tested on an external clinical dataset. The dataset included 2247 chest radiographs comprising CXRs from 1046 COVID-19 positive patients (positive on RT-PCR) and 1201 COVID-19 negative patients.

**Results:** We compared the performance of the model with three different radiologists by adjusting the model’s sensitivity as per the individual radiologist. The area under the curve (AUC) on the receiver operating characteristic (ROC) of the model was 0.87 [95% CI: 0.85, 0.89].

**Conclusion:** When compared to the performance of three expert readers, DxCOVID matched the output of two of the three readers. Disease-specific deep learning models using current technology are mature enough to match radiologists’ performance and can be a suitable tool to be incorporated into imaging workflows.

## Introduction

The world has seen multiple pandemics over the previous two millennia, but most of them occurred in the last two centuries. These included diseases like plague, Spanish flu, Asian Flu, Hong Kong flu, HIV, Swine flu, and Severe Acute Respiratory Syndrome (SARS) (1). Like most previous pandemics, Coronavirus Disease 2019 (COVID-19) primarily spreads via the respiratory route. The defining characteristics of a pandemic include widespread geographical extension, movement across territories, minimal pre-existing population immunity, and the novelty of genetic makeup associated with a high attack rate (2). While most of the factors have changed only a little over centuries, movement across continents has become very easy in the last century, allowing the disease to spread across the globe before being detected. According to World Health Organization (WHO) data from 2018, the availability of doctors in a developed country like Germany was approximately 43 per 10,000 people, whereas in the United States of America it was 23 per 10,000 people. The corresponding figures for developing nations like India (9.3 doctors per 10000 people) were significantly lower. In both cases, the numbers are insufficient to treat the general population, let alone provide adequate health care in the middle of a pandemic (3). In 2011, radiologists comprised only 3.7% of the total physician workforce in a developed nation like the USA, while in 2016 the number of radiographs performed globally had steadily increased to approximately 3.6 billion per annum. COVID-19 has only caused a further rise in the numbers (4,5).

The overall demand for chest radiography and Computed Tomography (CT) has increased as these imaging modalities have become a part of the COVID-19 diagnosis and management guidelines, as well as WHO recommendations because they allow the triage of the disease. While chest radiography is less sensitive as compared to CT, it is also less resource-intensive, and patients are exposed to lower doses of radiation. Moreover, in intensive care units, chest radiography is a valuable diagnostic tool to monitor disease progression. As of February 09, 2022, the total number of COVID-19 cases during the pandemic stood at over 401 million, with a mortality rate of nearly 5.7 million people globally (6). With the explosive increase in the population affected by COVID-19 and the number of radiographic studies performed, the turn-around time for radiology reports can be a bottleneck in the quick triage of studies that require urgent attention. Using artificial intelligence to perform quick triage can ensure study list reprioritization, ensuring prompt attention to urgent cases.

In the age of Computed Tomography (CT), our focus has been on developing a tool to assist detection of COVID-19 from chest radiographs because of the volume of chest radiographs performed daily and the emphasis placed on them for screening purposes. Even in Western nations with easy access to CT, chest radiography accounts for 25% of the total diagnostic procedures (7). Globally, these figures are even higher since chest radiography account for approximately 40% of all diagnostic procedures (3.6 billion) performed annually (8). The insight gained from an accurate interpretation of chest radiographs in COVID-19 can significantly alter the treatment plan advised by the attending physician and often obviate the need for CT. Even in Western nations, a chest radiograph is the first procedure to be advised for the purpose of screening. According to UNSCEAR, CT accounts for 8% of all radiological examinations and contributes to 47% of the total radiation dose in Level-I countries (9).

In this study, we present DxCOVID, a deep learning model for detecting COVID-19 from chest radiographs. We also compare the performance of DxCOVID to the interpretations by three board-certified radiologists.

## Material and Methods

### Data acquisition

This retrospective study was approved by the Institutional Review Board of our hospital. The data was anonymized, and explicit written informed consent was waived in view of the retrospective nature of the data collection and usage of completely anonymized patient datasets. The data was a mixed dataset of inpatient and outpatients collected from February 2021 to April 2021. This included 2247 patients, out of which 1046 were COVID-19 positive on RT-PCR, and the rest 1201 tested negative for COVID-19. The radiographs with negative COVID-19 RT-PCR reports were either normal or had other non-COVID-19 pathologies. Some patients required multiple radiograph examinations as their disease progressed. However, we avoided using multiple radiographs of the same patient as there is uncertainty regarding the performance evaluation of the deep learning model in such a case. To remove any bias in such a situation, a single radiograph per person was selected randomly. Hence, each radiograph corresponded to a single patient. De-identified chest radiographs that were well-exposed, well-centered, and covered the complete field of interest were included in the study. Poor quality radiographs that were end-expiratory, oblique, underexposed, overexposed, or exhibited motion blur and extraneous artifacts were excluded from the study. The methodology utilized for data collection, segregation, and analysis has been depicted in Fig 1.

**Figure 1:**
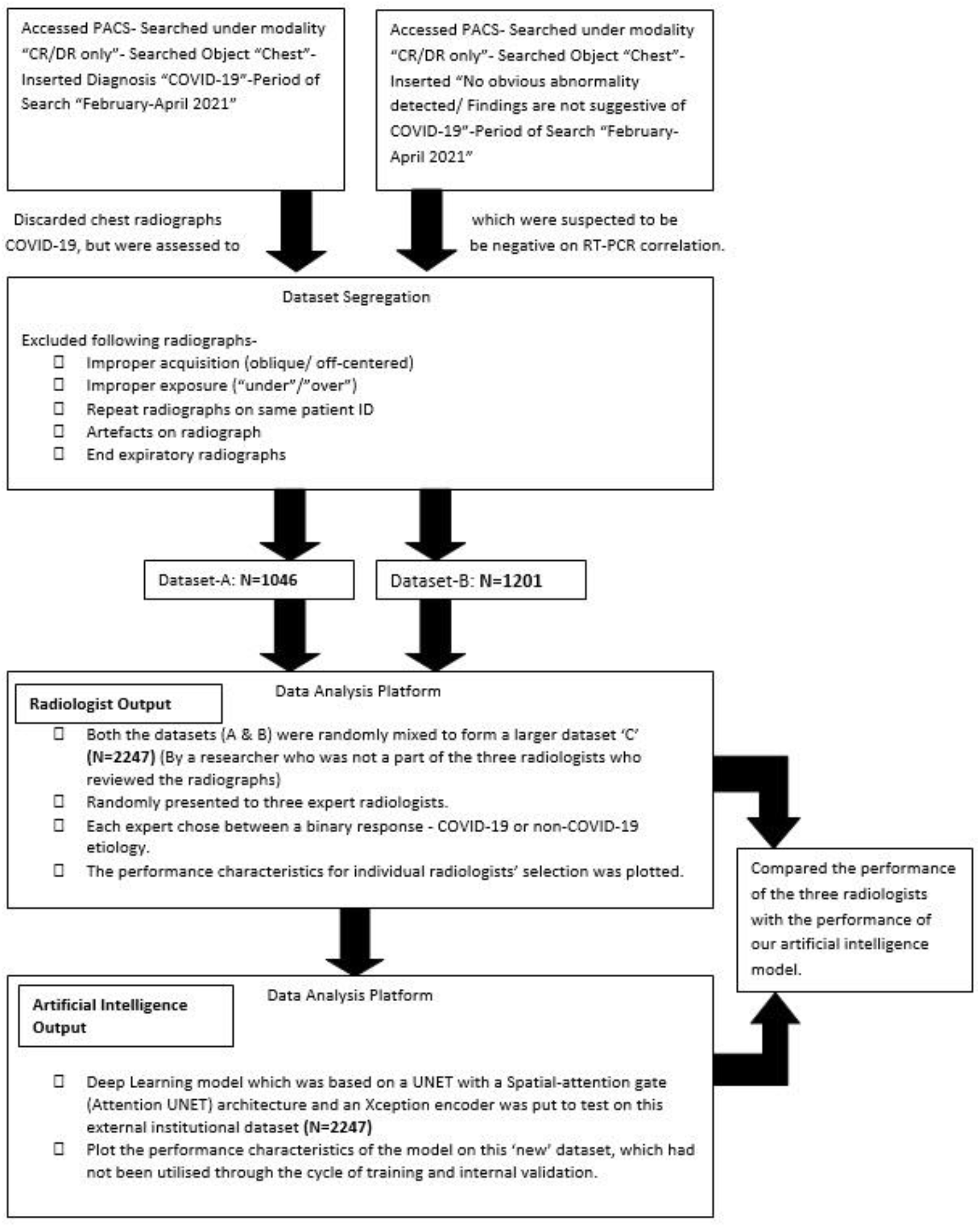
The detailed methodology used to acquire the external validation dataset has been elaborated. The method encompassed entering the appropriate query to the search system and extracting the resultant data from PACS (the hospital stores and accesses data using MedSynapse PACS). The data was segregated and finally uploaded onto a central data-analysis platform developed by DeepTek Medical Imaging Pvt Ltd in a Health Insurance Portability and Accountability Act (HIPAA) compliant manner. PACS: Picture Archiving and Communication System, CR: Computed Radiography, DR: Digital Radiography.

### Artificial Intelligence (AI) algorithm

We used a multistep process involving cycles of training and retraining before further validating or testing the AI model. The process involved data curation, anonymization, and pre-processing. The architecture used to train the AI model was enhanced UNET with a Spatial-attention gate (Attention UNET) and an Xception encoder. The processing step entailed checking for multiple parameters like validation loss and optimizing it based on the feedback. As a result, each cycle resulted in the development of a model that was trained and then modified based on the feedback from validation. Post model optimization, the model was evaluated on the external institutional hold-out dataset. This was done to minimize the overfitting of data. An illustration of the entire pipeline is summarised in Fig 2.

**Figure 2:**
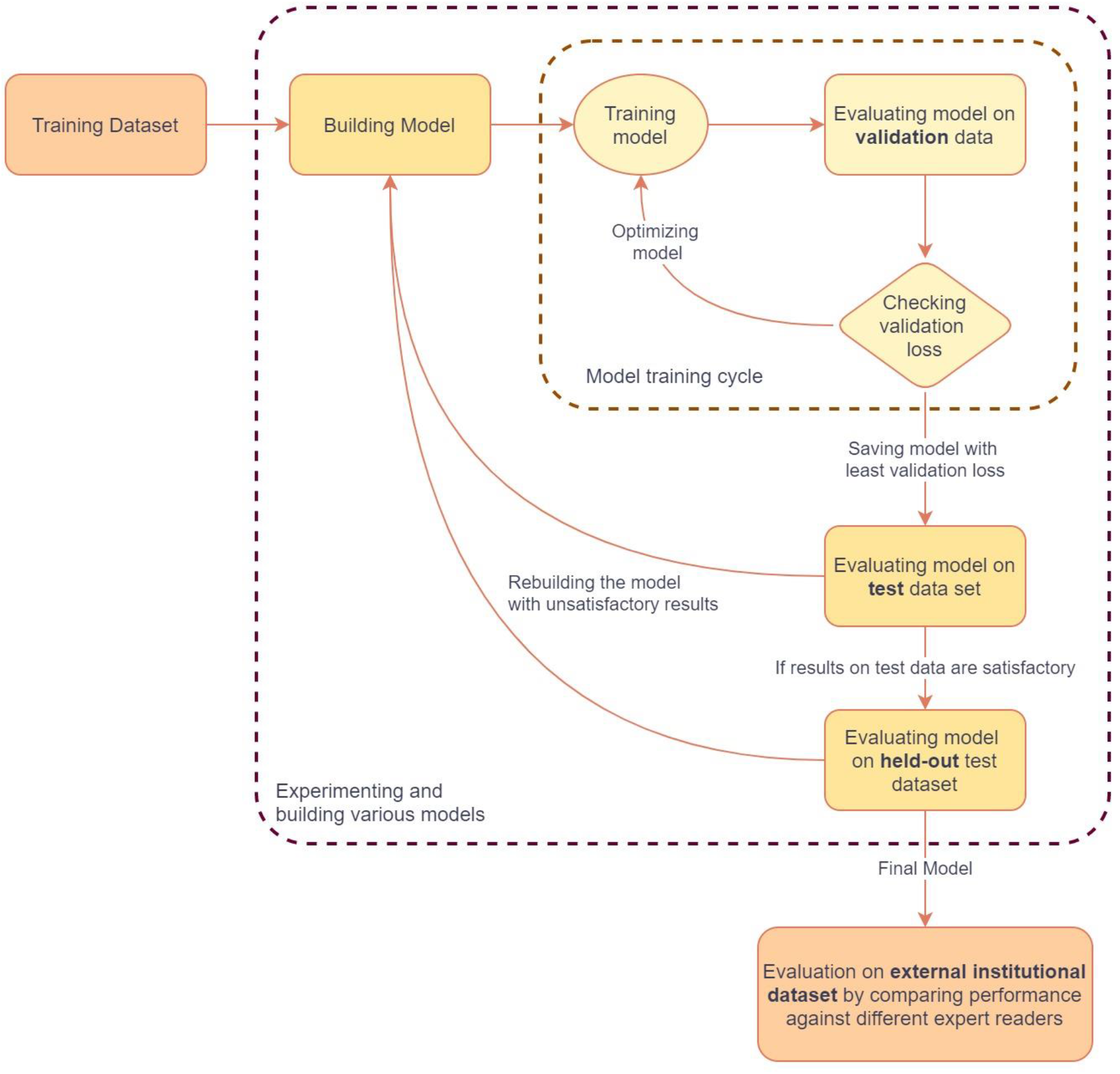
A workflow depicting the systematic development of the COVID-19 detection algorithm. It starts from the training dataset and subsequently goes through a rigorous and repetitive process of training and validation. The optimized model was evaluated on the external institutional dataset.

### Panel of radiologists for comparison

A panel of three board-certified radiologists read the entire dataset and labeled each image as COVID-19 positive or negative, individually. The panel included general radiologists with over 20-years of experience (R1), 23-years of experience (R2), and 27-years of experience (R3). The heterogeneous group of readers was chosen to ensure that the models’ output was tested against results from radiologists with different durations of work experience. Radiologists provided a binary output for each radiograph i.e., the radiograph was either marked positive for the presence of COVID-19 pneumonia pattern or negative for the given set of findings (focal consolidation, multifocal consolidations in a pattern and distribution typical for COVID-19).

### Statistical analysis

The 95% Confidence Interval (CI) was calculated using an empirical bootstrapping method. To evaluate the diagnostic performance of DxCOVID, the specificity, sensitivity, and accuracy were calculated in the entire test cohort of 2246 radiographs. The area under the receiver operating characteristic curve was calculated to assess the performance of the model.

## Results

### Model output

The output of the model contained a pixel-wise probability for the input image. The top 1% of predicted pixels were averaged out to obtain an overall probability of COVID-19. These probability scores were used to calculate the performance of the model. An illustration of the same is represented in Fig 3.

**Figure 3:**
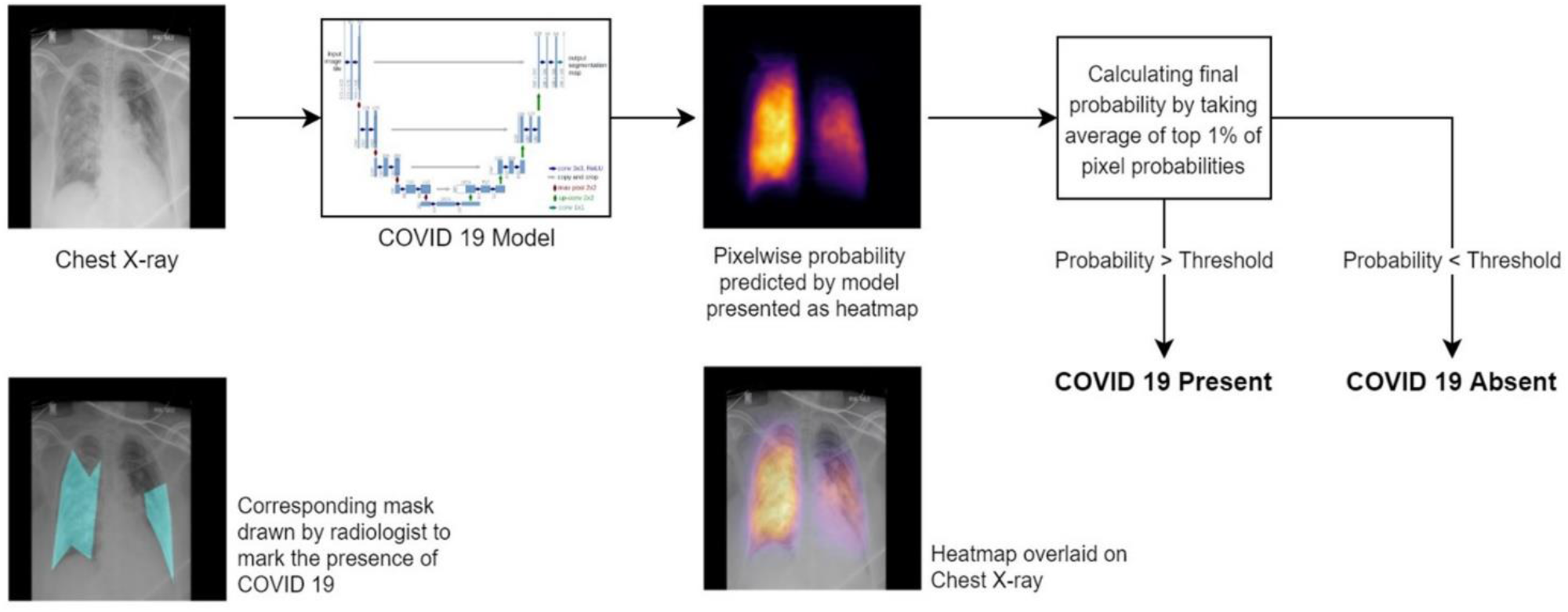
An illustration to depict the intrinsic working of the COVID-19 algorithm. The chest radiograph passes through multiple layers of the convolutional neural network and the output of the model is depicted using a heatmap. To simplify the process of assessment of the chest radiograph, the heatmap is superimposed on the chest radiograph. The final binary output of the presence or absence of COVID-19 was obtained by averaging out the top 1% of the pixel probabilities.

The possible localization of the manifestation of COVID-19 pneumonia is represented as a heat map by the model. The intense yellow color represents the high confidence score from which the model takes the decision to classify the area as COVID-19 positive or negative. To simplify the process of assessment of the chest radiograph, heatmaps were superimposed on chest radiographs (Fig 4).

**Figure 4:**
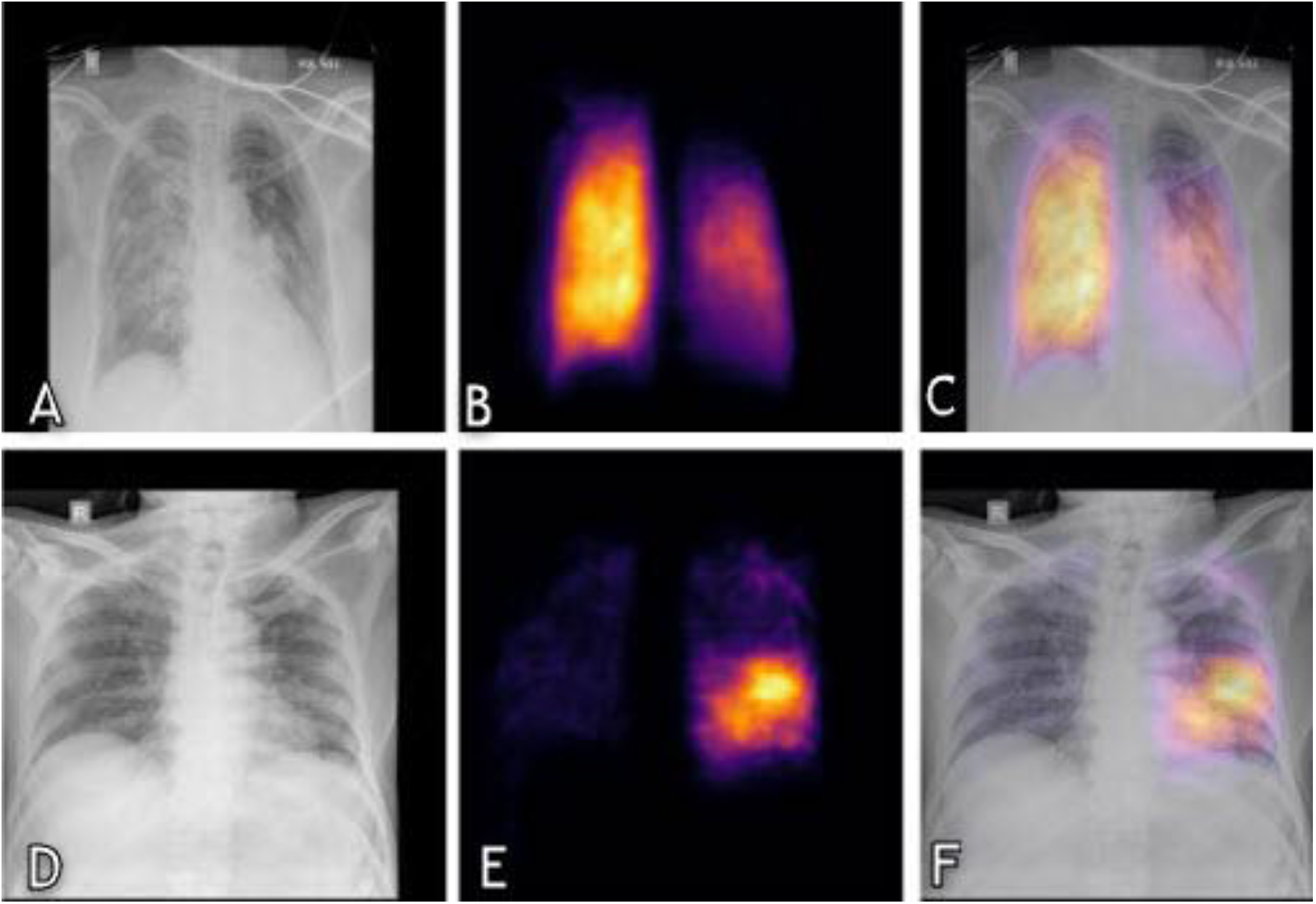
The panel of images comprises input chest radiographs, AP view (A, D). Once these chest radiographs were analyzed by the U-Net-based AI system, the system generated the heatmaps as depicted in corresponding images B, E. The final output (C, F) is also shown as an overlay of the AI model localizing the pathology on the chest radiograph, which helps the clinician in identifying suspicious regions affected with COVID-19 pneumonia.

### Model performance

The trained AI model was evaluated on the test dataset. Table 1 summarises the performance of DxCOVID (sensitivity, specificity, and accuracy) with its sensitivity adjusted to individual expert readers. The receiver operating characteristic (ROC) curve was used to evaluate the diagnostic accuracy of the model (Fig 5).

**Table 1:** Performance of the COVID-19 detection model with sensitivity adjusted to match that of the expert radiologists.

|  | **Expert Reader R1** | **DxCOVID** |
| --- | --- | --- |
| **Sensitivity** | 0.75 [0.72, 0.78] | 0.75 [0.72, 0.78] |
| **Specificity** | 0.81 [0.79, 0.83] | 0.82 [0.80, 0.84] |
| **Accuracy** | 0.79 [0.78, 0.81] | 0.80 [0.78, 0.81] |
|  | **Expert Reader R2** | **DxCOVID** |
| **Sensitivity** | 0.49 [0.45, 0.52] | 0.49 [0.45, 0.52] |
| **Specificity** | 0.96 [0.95, 0.97] | 0.94 [0.93, 0.95] |
| **Accuracy** | 0.81 [0.8, 0.83] | 0.80 [0.79, 0.82] |
|  | **Expert Reader R3** | **DxCOVID** |
| **Sensitivity** | 0.77 [0.74, 0.8] | 0.77 [0.74, 0.8] |
| **Specificity** | 0.86 [0.84, 0.88] | 0.80 [0.78, 0.82] |
| **Accuracy** | 0.83 [0.82, 0.85] | 0.79 [0.77, 0.81] |

**Figure 5:**
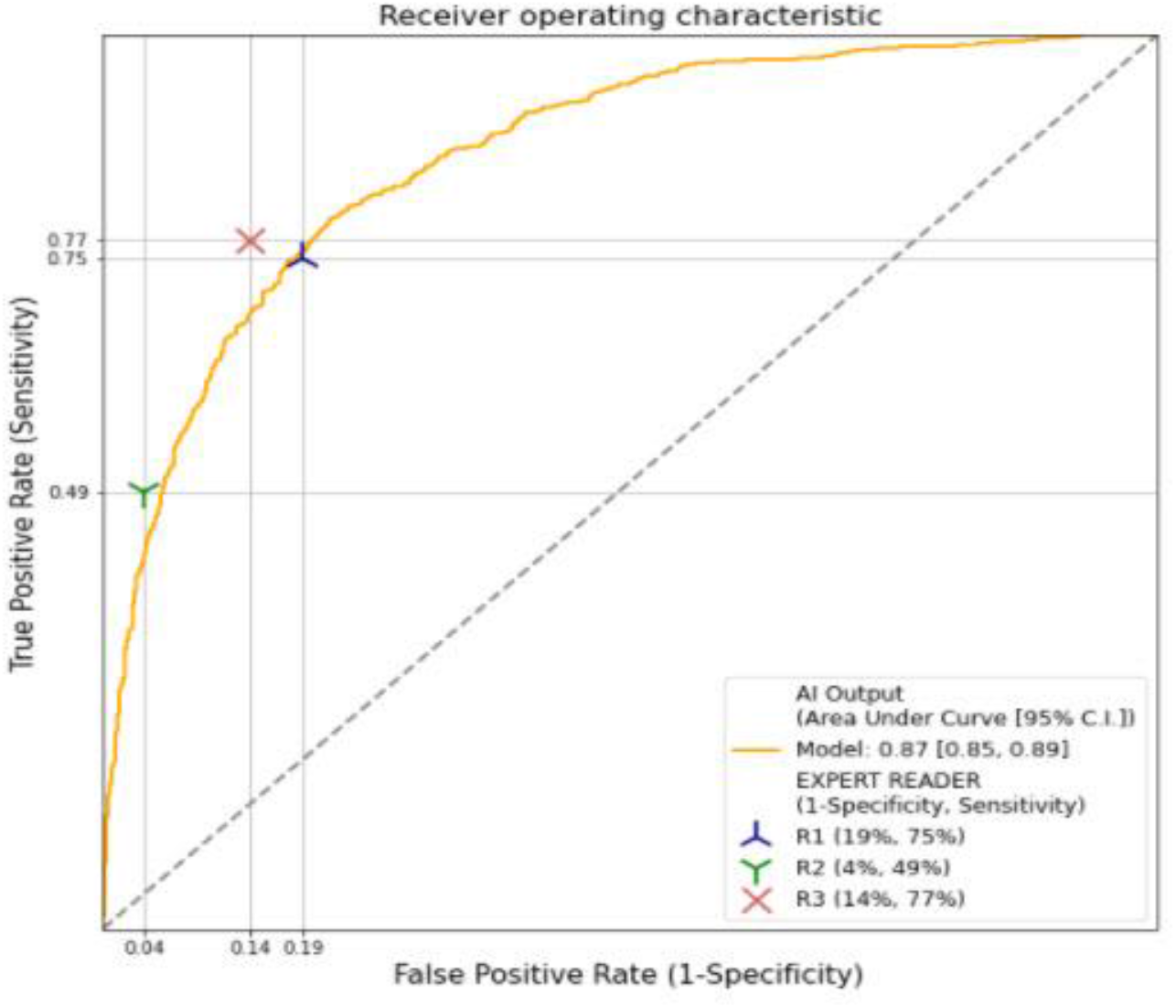
The receiver operator curve plots the output (AUC) of the three reviewing radiologists alongside the output of the externally validated AI model.

### Comparison of AI model with radiologists

The study design consisted of a multiparity assessment involving three board-certified radiologists (R1, R2, and R3). All readers were blinded to patient information and were instructed to read the radiographs independently and label them as positive or negative for COVID-19. Radiologists were trained to flag studies as suspicious for COVID-19 if the radiographs showed indications of either focal consolidation, multifocal consolidations in a pattern, and distribution typical for COVID-19, i.e., ground-glass opacity/consolidation in a predominantly peripheral distribution.

We compared the performance of DxCOVID with all three radiologists. As shown in Table 1, the expert reader (R1) exhibited a sensitivity of 0.75 [95% CI: 0.72, 0.78], specificity of 0.81 [95% CI: 0.79, 0.83], and an accuracy of 0.79 [95% CI: 0.78, 0.81]. The second reader (R2) had low sensitivity, i.e., 0.49 [95% CI: 0.45, 0.52], but extremely high specificity i.e., 0.96 [95% CI: 0.95, 0.97] for detecting COVID-19 pneumonia. The third reader (R3) had highest sensitivity i.e., 0.77 [95% CI: 0.74, 0.8], and accuracy i.e., 0.83 [95% CI: 0.82, 0.85] in detecting COVID-19 pneumonia. When compared to the performance of the three expert readers, the DxCOVID model matched the output of two of the three readers (R1 and R2 with the difference being statistically insignificant). The ROC results for all three expert readers and DxCOVID model are depicted in Figure 5. The model achieved the area under the curve of 0.87 [95% CI: 0.85, 0.89].

## Discussion

The Coronavirus Disease-2019 was declared as a Public Health Emergency of International Concern on 30 January 2020. A month later, on 11 March 2020, the WHO officially elevated its severity from an outbreak to a global pandemic, as multiple cases were being reported simultaneously from countries spanning multiple continents (10,11). Epidemiologists, researchers, physicians, and pharmaceutical companies backed by the government and/or privately funded organizations pooled resources to assess and develop tools to diagnose the disease and subsequently concentrate their efforts on the treatment aspect. The outcome was the development of reverse transcriptase-polymerase chain reaction (RT-PCR) kits to diagnose COVID-19. An array of blood panels including total leucocyte counts, ferritin, erythrocyte sedimentation rate, C-reactive protein (CRP), lactate dehydrogenase (LDH), and D-dimer were performed in all these cases, and while they could be linked to disease severity to varying extents, utilizing them for diagnosing the disease proved difficult (12,13). Researchers explored the usage of time-tested imaging modalities, such as chest radiography and computed tomography (CT) for diagnostic purposes and subsequently documented the specific signs on imaging for COVID-19 (14,15).

A detailed framework for the role of imaging in diagnosing COVID-19 was published by the Fleischner Society (16). The document included elaborate algorithms depicting when and how to utilize imaging techniques in COVID-19 and interpret them concurrently with other investigations. With lockdown imposed in most of the countries alongside a shortage of beds and medical equipment, patients with mild symptoms, particularly in the developing nations, received treatment at home, while patients with more advanced symptoms were rushed to emergency departments. In this subset of patients, radiography had the advantage of being portable, quick, low in radiation exposure, and an adjunct tool allowing the monitoring of disease progression (17).

While pneumonia and bronchopneumonia have characteristic imaging features, COVID-19 pneumonia has a wide range of imaging manifestations. COVID-19 radiographs have proven to be difficult to read, particularly in patients with a mild form of the disease. This is because the COVID-19 pneumonia pattern is similar to the pattern manifested in other infectious diseases, such as pulmonary thromboembolism and pulmonary hemorrhage (18).

Extensive research in artificial intelligence and radiology has resulted in some actual tested and working algorithms capable of detecting COVID-19. A study conducted by researchers from the North-Western Memorial Health Care Center on a dataset of 5853 patients resulted in the development of the DeepCOVID-XR algorithm. While their training and validation sets were among the largest to date, the actual subset of data where comparisons to board-certified radiologists were made was only 300 (19). In our case, the results of all 2247 patient datasets were used to test the model were compare its performance with three board-certified radiologists.

Zhang et al. developed a deep learning model called CV-19 that utilized a dataset of 2020 COVID-19 and 3148 non-COVID-19 patients. They utilized the test dataset of 500 radiographs to ascertain the system’s threshold that produced an AUC of 0.96, tested against the outputs of three radiologists^20^. Herein, we tested the model DxCOVID in a similar rigorous manner, and the output of the AI model was plotted against the output of three experts. In our study, we avoided utilizing multiple radiographs of the same patient and instead selected a single study for each patient so that 2247 chest radiographs equated to 2247 patient datasets. By doing so, we avoided the debate of whether utilizing multiple radiographs of the same patient could lead to a change in performance evaluation, a concern voiced by Zhang et al. (20).

In a recent study by Roberts et al. (21), the researchers have evaluated the suitability of existing models, their clinical relevance, the pitfalls of the various algorithms, and their findings. The researchers concluded that while various AI models are available, none of the studies produced a clinically relevant model, and therefore, the results are not generalizable to the masses. The document reveals that, while the focus is on developing COVID-19 AI models in response to the pandemic, the actual utility is still highly debatable in real-world practices. To that end, they have identified several potential causes and have also recommended guidelines to be followed while developing and testing the algorithms for enhanced comparisons. We took cues from this study, and our model outputs were calibrated based on the individual radiologists’ sensitivity, rather than a separate ground truth or RT-PCR reports.

Our study has some limitations. First, the external test set was sourced from a single institution. It is critical to test the AI algorithm across multiple datasets distributed across geographical regions to ensure that the model’s results are generalizable across cohorts and geographies. Second, we utilized RT-PCR as the gold standard for the diagnosis of COVID-19 infections. However, RT-PCR has a limited sensitivity of approximately 71%, so there may be cases where the person is COVID-19 positive on chest radiographs but negative on RT-PCR results. Third, our study does not incorporate clinical parameters and does not attempt at categorising patients based on COVID-19 severity scores. We avoided providing COVID-19 scores based on chest radiographs as unlike chest CT scans, there is often interobserver disagreement on the extent of lung involvement in radiography for reasons encompassing different acquisition protocols, image quality, and radiologist opinion. The development of an AI system based on the consensus scoring of a few radiologists from an isolated geographical location may not represent the consensus of radiologists globally. However, some researchers did attempt to build such a system, like the one developed by Borghesi & Maroldi (22) on a small dataset of 100 patients. Attempts were also made by Monaco et al. (23) with the dataset of 295 patients and Orsi et al. (24) with the dataset of 155 patients to produce scoring systems for chest radiographs and link them with certain key clinical and laboratory parameters. However, their results were limited by factors from variable interobserver disagreements ranging from moderate to severe, a limited dataset, datasets confined to narrow geographical locations, and the consensus scoring systems were developed by a very limited number of radiologists.

In conclusion, we developed a DL model (DxCOVID) for detecting COVID-19 patterns from digital chest radiographs. This model achieved an area under the curve (AUC) of 0.87 [95% CI: 0.85, 0.89] when tested on the institutional dataset of 2247 radiographs from a single institution. When compared to the performance of the three expert readers, the DxCOVID model matched the output of two of the three readers (R1 and R2). AI could serve as a reliable second reader, augmenting the result interpretation by experts. Directions for future research include the development of a successfully deployable model that incorporates various physical and clinical parameters in addition to the imaging findings from multi-institutional datasets and provide a combined output that can adequately prognosticate the patient.

## Data Availability

All data produced in the present study are available upon reasonable request to the authors

## References

1. Huremović D. Brief history of pandemics (pandemics throughout history). In Psychiatry of pandemics 2019 (7–35). Springer, Cham.

2. Morens DM, Folkers GK, & Fauci AS. What is a pandemic?. J Infect Dis 2009; 200; 1018–1021.

3. http://www.who.int/data/gho/data/indicators/indicator-details/GHO/medical-doctors-(per-10-000-population) [Accessed on 1st September, 2021]

4. Rosenkrantz AB, Hughes DR, Duszak Jr R. The US radiologist workforce: an analysis of temporal and geographic variation by using large national datasets. Radiology; 2016; 279; 175–184.

5. https://www.who.int/news-room/feature-stories/detail/to-x-ray-or-not-to-x-ray- [Accessed on 1st September, 2021]

6. https://www.worldometers.info/coronavirus/ [Accessed on February 9, 2022]

7. Speets AM, van der Graaf Y, Hoes AW, et al. Chest radiography in general practice: indications, diagnostic yield and consequences for patient management. Br J Gen Pract 2006; 56; 574–578.

8. Bray F, Ferlay J, Soerjomataram I, et al. Global cancer statistics 2018: GLOBOCAN estimates of incidence and mortality worldwide for 36 cancers in 185 countries. CA Cancer J Clin 2018; 68; 394–424. Erratum in: CA Cancer J Clin 2020; 70; 313.

9. Sun Z, Ng KH. Use of radiation in medicine in the Asia-Pacific region. Singapore Med J. 2012; 53; 784–788.

10. Cucinotta D, Vanelli M. WHO declares COVID-19 a pandemic. Acta Biomed 2020; 91; 157.

11. Bernheim A, Mei X, Huang M, et al. Chest CT findings in coronavirus disease-19 (COVID-19): relationship to duration of infection. Radiology 2020; 20; 200463.

12. https://www.who.int/teams/blueprint/covid-19 [Accessed on 10th August, 2021]

13. Stegeman I, Ochodo EA, Guleid F, et al. Cochrane COVID-19 Diagnostic Test Accuracy Group. Routine laboratory testing to determine if a patient has COVID-19. Cochrane Database Syst Rev 2020; 11: CD013787.

14. Fang Y, Zhang H, Xie J, et al. Sensitivity of chest CT for COVID-19: comparison to RT-PCR. Radiology 2020; 296; e115–117.

15. Lyu P, Liu X, Zhang R, et al. The performance of chest CT in evaluating the clinical severity of COVID-19 pneumonia: identifying critical cases based on CT characteristics. Invest Radiol 2020; 55; 412–421.

16. Rubin GD, Ryerson CJ, Haramati LB, et al. The role of chest imaging in patient management during the COVID-19 pandemic: a multinational consensus statement from the Fleischner Society. Radiology 2020; 296; 172–180.

17. Eisenhuber E, Schaefer-Prokop CM, Prosch H, et al. Bedside chest radiography. Respir Care 2012; 57; 427–443.

18. Duzgun SA, Durhan G, Demirkazik FB, et al. COVID-19 pneumonia: the great radiological mimicker. Insights Imaging 2020; 11; 1–5.

19. Wehbe RM, Sheng J, Dutta S, et al. DeepCOVID-XR: an artificial intelligence algorithm to detect COVID-19 on chest radiographs trained and tested on a large US clinical data set. Radiology 2021; 299; e167–176.

20. Zhang R, Tie X, Qi Z, et al. Diagnosis of coronavirus disease 2019 pneumonia by using chest radiography: Value of artificial intelligence. Radiology 2021; 298 ; e88–97.

21. Roberts M, Driggs D, Thorpe M, et al. Common pitfalls and recommendations for using machine learning to detect and prognosticate for COVID-19 using chest radiographs and CT scans. Nat Mach Intell 2021; 3; 199–217.

22. Borghesi A, Maroldi R. COVID-19 outbreak in Italy: experimental chest X-ray scoring system for quantifying and monitoring disease progression. Radiol Med 2020; 125; 509–513.

23. Monaco CG, Zaottini F, Schiaffino S, et al. Chest x-ray severity score in COVID-19 patients on emergency department admission: a two-centre study. Eur. Radiol. Exp 2020; 4; 1–7.

24. Orsi MA, Oliva G, Toluian T, Pittino CV, Panzeri M, Cellina M. Feasibility, reproducibility, and clinical validity of a quantitative chest X-ray assessment for COVID-19. Am J Trop Med Hyg 2020; 103; 822.

